# The concentration of several perfluoroalkyl acids in serum appears to be reduced by dietary fiber

**DOI:** 10.1101/2020.07.15.20154922

**Authors:** Michael W. Dzierlenga, Debra R. Keast, Matthew P. Longnecker

## Abstract

Fiber-rich food intake has been associated with lower serum concentrations of perfluoroalkyl substances (PFAS) in some studies and dietary fiber was related to lower serum PFAS in a recent study. Given the previous epidemiologic data suggesting that fiber might decrease serum PFAS concentrations, we examined the relation of serum PFAS concentrations to intake of dietary fiber in National Health and Nutrition Examination Survey (NHANES) data. We examined the PFAS-fiber association among 6,482 adults who participated in the NHANES, 2005-2016. Fiber intake was estimated based on two 24-hour diet recalls. We adjusted the models for determinants of PFAS and potentially confounding factors such as intake of foods reported to increase PFAS exposure. Results were expressed as the percent difference in PFAS concentration per interquartile range (IQR) increase in fiber (and 95 percent confidence interval), and the NHANES sampling parameters were used to make the results generalizable to the U.S. The adjusted percent difference in perfluorooctanoic acid (PFOA) per IQR increase in fiber was −3.64 (−6.15, −1.07); for perfluorooctane sulfonic acid (PFOS) was −6.69 (−9.57, −3.73), and for perfluorononanoic acid (PFNA) was −8.36 (−11.33, −5.29). These results suggest that dietary fiber increases the gastrointestinal excretion of PFOA, PFOS, and PFNA. Because fiber also lowers serum cholesterol, in some studies of the serum cholesterol-PFAS relationship confounding by fiber may be worth evaluating.

## Introduction

Perfluorinated alkyl substances (PFAS) have been associated with a number of health outcomes in humans, even when the exposure was below that found to have an effect in animals (EPA 2016a; EPA 2016b; EFSA 2018). A prime example is increased serum cholesterol (Steenland et al., 2009). In a recent report based on the National Health and Nutrition Examination Survey (NHANES), the cholesterol-PFAS association was 1.5 mg/dl per ng/ml of perfluorooctanoic acid (PFOA) in serum (Dong et al., 2019). Confounding is a plausible explanation for an association of such modest size. Given the paucity of animal data supporting adverse effects of PFAS on cholesterol at background levels of exposure, a natural question to ask was: could there be an as-yet-unidentified confounding factor that might explain this relationship? We identified consumption of dietary fiber as a possible source of confounding, as it has a well-known inverse association with cholesterol and may decrease PFAS levels through increases in gastrointestinal excretion.

Dietary fiber decreases absorption of several constituents of food such as cholesterol, fat, and iron (Grube et al., 2013; Hallberg and Hulthén, 2000; Jesch and Carr, 2017). Fiber also decreases the absorption of bile acids which contributes to its serum-cholesterol-lowering effect (Brown et al., 1999; Ellegåard and Andersson, 2007). Dietary fiber decreases absorption by adsorbing substances or trapping them in viscous gel matrix (Naumann et al., 2019). Fiber-rich food intake has been associated with lower serum PFAS concentration in some studies (Halldorsson et al., 2008; Skuladottir et al., 2015) and dietary fiber was related to lower serum PFAS in a recent study (Lin et al., 2020: Supplementary Material Figure S4).

PFAS were used or have ongoing use in manufacturing of a variety of products, including consumer-use products (Buck et al., 2011; Interstate Technology Regulatory Council (ITRC), 2020). Almost everyone in developed countries has detectable amounts of PFAS in their serum, due to exposure via contaminated food and other sources (Sunderland et al., 2019). The association of human health outcomes with PFAS have been, in some cases, used as the critical endpoint in risk assessments, even when the epidemiologic results were from populations with background-level exposure (EFSA CONTAM Panel, 2020, 2018).

Given the previous epidemiologic data suggesting that fiber might decrease serum PFAS concentrations and the possibility that a PFAS-fiber association might confound the results of studies on health outcomes and PFAS, we examined the relation of serum PFAS concentrations to intake of dietary fiber in NHANES data. The NHANES now has more than 10 years’ worth of data on serum PFAS concentrations among subjects with two 24-hour diet recalls (Centers for Disease Control and Prevention (CDC) National Center for Health Statistics (NCHS), 2015).

## Methods

### Study population

Data on NHANES participants from the 2005-2006 through 2015-2016 waves were used. Because the food frequency questions about fish consumption were asked only of women of reproductive age in 2003-2004, that NHANES wave was not included. (Earlier waves had only one 24-hour diet recall per subject.) Participants were included in the main analysis if they had complete dietary data, measurement of PFAS levels, and data for all other covariates (see *other covariates* below). Participants who reported using bile acid sequestrants were excluded as these drugs may interfere with absorption of PFAS from the gut (Genuis et al., 2010). Participants were also excluded if they were under 20 or over 79 years old, if they were pregnant, if they were breastfeeding, or if they were undergoing dialysis.

### PFAS Levels

PFOA, perfluorosulfonic acid (PFOS), and perfluorononanoic acid (PFNA) were measured in all the waves from 2005-2006 through 2015-2016. We selected PFOA, PFOS and PFNA as the PFAS to examine based on the high proportion of participants with detectable levels of those compounds in human serum (>99%) and the tendency for the levels of those chemicals to be associated with cholesterol levels in serum (Dong et al., 2019; Geiger et al., 2014; Nelson et al., 2010; Steenland et al., 2009). PFAS were quantified in a subsample of one third of the NHANES participants. For waves from 2013-2014 and 2015-2016, in which the linear and branched isomers of PFOA and PFOS were measured separately, we used the sum of the linear and branched isomers. As mentioned previously, the three chemicals were measured at levels above the limit of detection (LOD) in most participants, with the exception of the branched isomer of PFOA, which was below the LOD in most of the participants in which the isomers of PFOA were separately measured. Measurements of levels below the LOD were imputed with LOD/sqrt(2). The sum of PFOA was mostly determined by the linear isomer, with a mean ratio of linear to branched PFOA of 25.8.

### Dietary Information

The dietary survey portion of NHANES consisted of two 24-hr diet recalls, and a targeted food frequency questionnaire covering fish and seafood consumption within the last 30 days. Based on the consumed amount of individual foods reported in the 24-hr recalls, nutrient intakes were derived using the NHANES wave-specific Food and Nutrient Database for Dietary Studies (Food Surveys Research Group, Agricultural Research Service, USDA, 2018). Total dietary fiber was the fiber remaining after a food’s digestion by α-amylase, protease, and amyloglucosidase (Lee et al., 1992).

Foods that have been reported to contain high levels of PFAS (Domingo et al., 2012; Ericson et al., 2008; Haug et al., 2010; Noorlander et al., 2011; Tittlemier et al., 2007; Vestergren and Cousins, 2013), or whose consumption has been associated with higher PFAS levels (Arrebola et al., 2018; Averina et al., 2018; Brantsaeter et al., 2013; Eriksen et al., 2011; Jain, 2014; Liu et al., 2017; Park et al., 2019; Skuladottir et al., 2015; Zhou et al., 2019) were considered as potentially confounding factors: fish and seafood, meat and meat products, milk and dairy products, eggs and egg products, and popcorn. For fish and seafood, adjustment was based on the number of reported times fish and seafood were eaten in the past 30 days. For the other food categories, adjustment was based on the grams/d of each food category consumed, averaged over the two 24-hr recalls. Details about the classification of food recall data into food categories are presented in the supplemental information (Table S1). Total energy intake was also considered as a covariate.

### Other Covariates

For the adjusted analyses, we adjusted a priori for sex, age, race and ethnicity (5 categories), NHANES wave, income to poverty ratio, and BMI (kg/m^2^). For women, we also considered adjustment for parity. We also considered adjustment for cigarette smoking (never [<100 cigarettes lifetime], former [does not currently smoke cigarettes], smoker [<1 pack per day average last 30 days], heavy smoker [≥1 pack per day average last 30 days]), and alcohol intake (never [<12 drinks lifetime], former [0 drinks last 12 mo.], light drinker [<1 drink per week last 12 mo.], drinker [<7 drinks per week last 12 mo.], heavy drinker [≥7 drinks per week last year).These covariates were chosen based on their possible influence on diet composition, energy intake, dietary fiber, or serum PFAS level (Figure S1).

### Statistical Methods

Univariate distributions were analyzed using median and quartiles for continuous variables and by percentage for categorical variables. In a preliminary step, energy-intake-adjusted fiber intake was calculated as the residual of the regression of dietary fiber on total caloric intake. Energy-intake-adjustment produces a variable which represents dietary intake from a relative and not absolute perspective (Willett et al., 1997). Energy-adjusted fiber residuals reflect variation in diet composition, whereas crude fiber intakes reflect variation in levels of energy intake and diet composition. The Pearson correlation of energy intake with crude fiber was 0.53. One advantage of the “residual approach” is that the IQR of the energy-adjusted residuals more readily corresponds to a realistic difference in diet than does an IQR difference in crude fiber. Fiber and caloric intake were log-transformed to normalize the distributions for the regression. As the residuals of the regression were centered at zero, the mean crude fiber intake was added back to the residuals to produce an easier to interpret metric. The energy-intake adjustment was done before averaging the two 24-hr dietary recalls. We fit a regression model of log_e_ transformed PFAS with energy-intake-adjusted dietary fiber intake as the key independent variable (continuous, original scale), with adjustment for the a priori and other factors considered. Dietary and lifestyle variables were removed in order of smallest impact of their removal on the β for PFAS-fiber, with the condition that variables were removed such that the modified β varied by less than 5% than the β from the full model. Linear and second order models for fiber were also tested, with the quadratic model selected if the AIC was improved (decreased) by 4 or greater. We also performed regressions with quartile of energy-intake-adjusted total dietary fiber intake as the independent variable.

We also examined effect modification by comparing models with and without interaction terms for dietary fiber consumption and each confounding variable in the full model. The comparison was performed using a decrement in AIC ≥4 indicating a better-fitting model. To see if the PFAS-fiber association varied by source of fiber we fit models with total fiber replaced by a group of variables representing fiber intake from specific types of food. Details about the food categories used for this analysis are in the supplemental information (Table S2). For comparison with the g/d fiber results, we fit similar models of g/d of food from the categories of fiber-containing foods.

In sensitivity analyses, we investigated the possibility of bias due to missing data using multiple imputation with chained equations (Buuren and Groothuis-Oudshoorn, 2011). We performed the analysis using fully conditional specification to impute the missing data for the adjustment factors. Five chains were used for imputation with 25 iterations performed for each chain. Convergence was evaluated by examining plots of variable mean and standard deviation across chains and iterations.

We also fit versions of our models that instead of using energy-adjusted fiber values (and energy intake for the full model), included crude fiber and energy intake. We also estimated the PFAS-fiber associations that would have been observed in the absence of measurement error in fiber intake, given certain assumptions. This is described in the supplemental information (Section 1).

All statistical analyses were carried out in R (R Core Team, 2018). In the initial diet calculations, the 2-day dietary sample weight was used for the 24-hr dietary recall data to account for day-of-the-week variation and other factors. In the regression models the sampling parameters and survey weights for the PFAS measurements were used. The R package “survey” was used to perform the regression (Lumley, 2019a, 2004). The R packages “mice” and “mitools” were used to perform and analyze the multiple imputation (Buuren and Groothuis-Oudshoorn, 2011; Lumley, 2019b).

## Results

After excluding subjects with incomplete information (Figure S2), data for 6,482 people were included in the analysis. Most exclusions (more than 1% of the total initial population) were due to age, lack of outcome measurement, or lack of one or both 24-hr diet recalls. The characteristics of the 6,482 reflected that of the U.S. adult population (Table 1). The median fiber intake was consistent with the reported U.S. mean of 16 g/d (Hoy and Goldman, 2014), recognizing the slight skewness of the distribution; the distribution of energy-adjusted fiber was slightly narrower than for the crude distribution, as expected. Popcorn consumption had 0 g/day for the 75^th^ quartile. Further analysis showed that 619 individuals (9.5%) had popcorn intake greater than 0 g/day.

**Table 1.** Population characteristics. Number (and percentage) are shown for categorical variables. Median (and quartiles) are shown for continuous variables.^a^

| Characteristic | Number or Median |
| --- | --- |
| Sample Size | 6482 |
| Sex |  |
| Female | 3260 (50.3%) |
| Male | 3222 (49.7%) |
| Age | 47 (34, 58) |
| Race/Ethnicity |  |
| Mexican American | 495 (7.6%) |
| Other Hispanic | 325 (5.0%) |
| Non-Hispanic White | 4603 (71.0%) |
| Non-Hispanic Black | 661 (10.2%) |
| Other Race | 398 (6.1%) |
| Income-Poverty Ratio | 3.2 (1.6, 5) |
| Education Level |  |
| Less than 9<sup>th</sup> Grade | 284 (4.4) |
| 9-11<sup>th</sup> Grade | 659 (10.2) |
| High School Grad./GED | 1435 (22.1) |
| Some College or AA Degree | 2048 (31.6) |
| College Graduate or Above | 2056 (31.7) |
| BMI (kg/m<sup>2</sup>) | 27.9 (24.3, 32.7) |
| Parity (among women) |  |
| 0 | 1587 (24.5%) |
| 1 | 1007 (15.5%) |
| 2 or more | 3889 (60.0%) |
| PFOA (ng/ml) | 2.8 (1.8, 4.5) |
| PFOS (ng/ml) | 9.1 (5.0, 15.9) |
| PFNA (ng/ml) | 0.9 (0.6, 1.4) |
| Smoking |  |
| Never [<100 lifetime cigarettes] | 3549 (54.8%) |
| Former [not current smoker] | 1591 (24.5%) |
| Smoker [< 1 pack per day] | 860 (13.3%) |
| Heavy Smoker [≥ 1 pack per day] | 481 (7.4%) |
| Alcohol Use |  |
| Never [<12 lifetime drinks] | 647 (10.0%) |
| Former [0 drinks last 12 months] | 977 (15.1%) |
| Light Drinker [<1 drink per week] | 2045 (31.5%) |
| Drinker [<7 drinks per week] | 1751 (27.0%) |
| Heavy Drinker [≥7 drinks per week] | 1063 (16.4%) |
| Dietary Intake |  |
| Crude Dietary Fiber (g/day)<sup>b</sup> | 15.4 (10.9, 21.8) |
| Energy Adjusted Fiber (g/day)<sup>b</sup> | 14.4 (10.8, 19.1) |
| Total Energy Intake (kcal/day) | 2000 (1563, 2588) |

<sup>a</sup> Sampling parameters were used in calculating the above values.
<sup>b</sup> Dietary fiber was log-transformed for energy adjustment. The geometric mean crude dietary fiber was 14.5 g/day and for energy-adjusted fiber was 14.2 g/day.

Energy-adjusted dietary fiber was correlated with all the other dietary intake variables, based on statistical significance (Table 2). In terms of the magnitude of the correlation, Fish & Shellfish had the largest correlation (positive), followed by Meat & Meat products (negative) and Popcorn (positive).

**Table 2.**
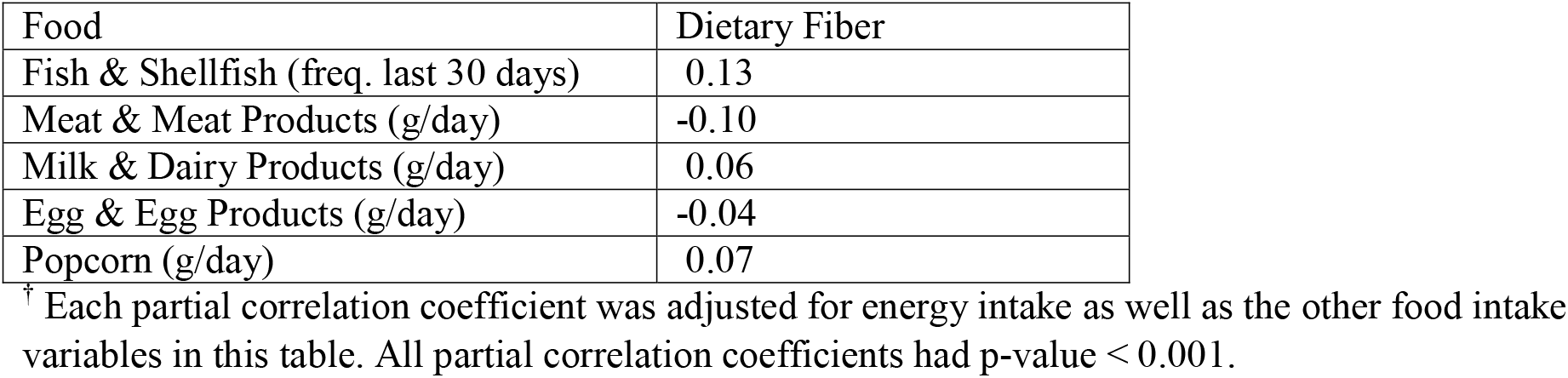
Partial correlation coefficients between dietary fiber and food intake variables.^†^

| Food | Dietary Fiber |
| --- | --- |
| Fish & Shellfish (freq. last 30 days) | 0.13 |
| Meat & Meat Products (g/day) | -0.10 |
| Milk & Dairy Products (g/day) | 0.06 |
| Egg & Egg Products (g/day) | -0.04 |
| Popcorn (g/day) | 0.07 |

<sup>†</sup> Each partial correlation coefficient was adjusted for energy intake as well as the other food intake variables in this table. All partial correlation coefficients had p-value < 0.001.

Popcorn is a source of dietary fiber, so a positive correlation was expected. Seafood and meat do not contain fiber, so the correlations with fiber must be due to differences in the overall diet of frequent seafood and meat consumers.

In both the unadjusted and adjusted models of lnPFAS, the percent difference per IQR increment in energy-adjusted dietary fiber was inverse for all three compounds (Table 3). Comparing the unadjusted and adjusted results, addition of potentially confounding variables decreased the magnitude of association for PFOA, slightly decreased it for PFOS, and increased the magnitude of the association for PFNA. In the adjusted analyses, PFNA had the strongest association, followed by PFOS and PFOA. Only the model of PFNA was improved by the addition of a quadratic term for energy-adjusted fiber. Eliminating covariates whose inclusion had a small effect on the β for dietary fiber resulted in in energy intake, dairy, eggs, income to poverty ratio and smoking being removed from the PFOA model, energy intake, meat, dairy, eggs, popcorn, income to poverty ratio, parity, wave^2^ and alcohol being removed from the PFOS model, and energy intake, dairy, eggs, popcorn, income to poverty ratio, parity and alcohol being removed from the PFNA model. The coefficients for all terms included in the final models are shown in Tables S3-S5.

**Table 3.**
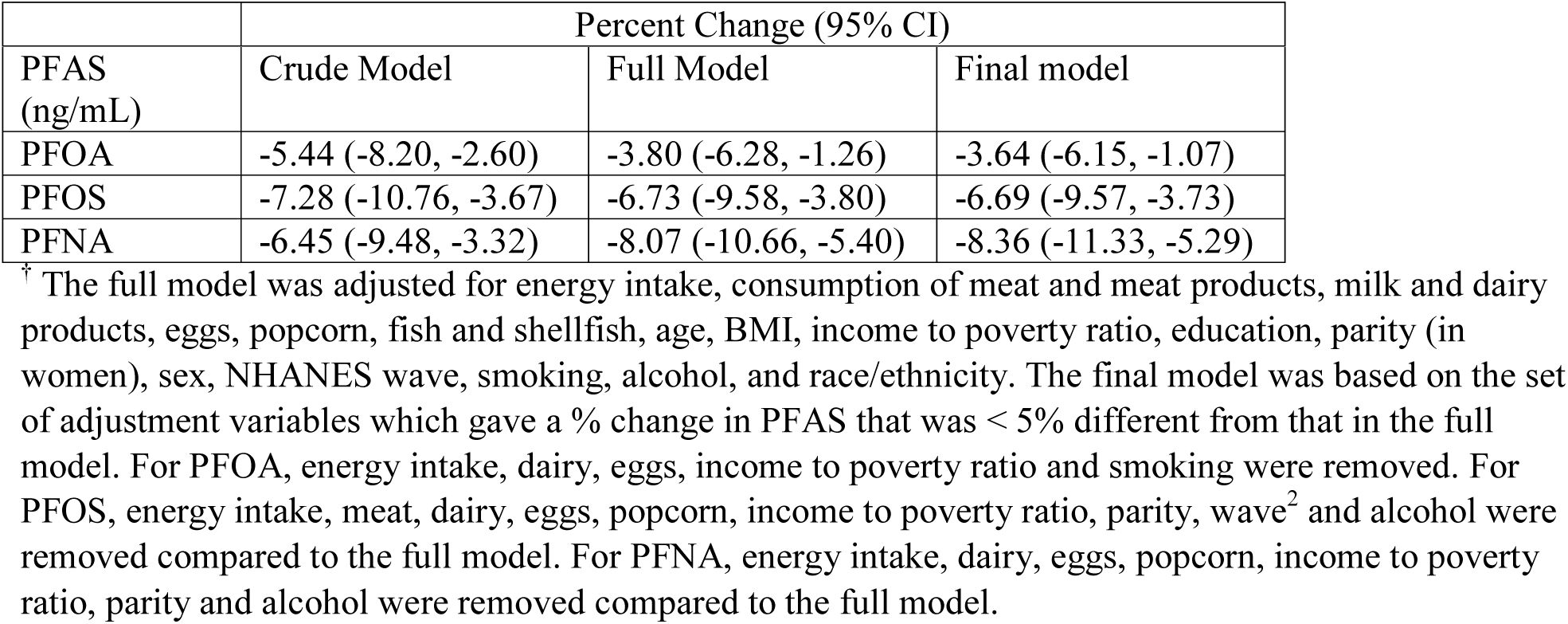
Percent change in PFAS level per interquartile distance (IQR) increase in energy-adjusted fiber intake using three different models.^†^

| PFAS<br>(ng/mL) | Crude Model | Full Model | Final model |
| --- | --- | --- | --- |
| PFOA | -5.44 (-8.20, -2.60) | -3.80 (-6.28, -1.26) | -3.64 (-6.15, -1.07) |
| PFOS | -7.28 (-10.76, -3.67) | -6.73 (-9.58, -3.80) | -6.69 (-9.57, -3.73) |
| PFNA | -6.45 (-9.48, -3.32) | -8.07 (-10.66, -5.40) | -8.36 (-11.33, -5.29) |

<sup>†</sup> The full model was adjusted for energy intake, consumption of meat and meat products, milk and dairy products, eggs, popcorn, fish and shellfish, age, BMI, income to poverty ratio, education, parity (in women), sex, NHANES wave, smoking, alcohol, and race/ethnicity. The final model was based on the set of adjustment variables which gave a % change in PFAS that was < 5% different from that in the full model. For PFOA, energy intake, dairy, eggs, income to poverty ratio and smoking were removed. For PFOS, energy intake, meat, dairy, eggs, popcorn, income to poverty ratio, parity, wave<sup>2</sup> and alcohol were removed compared to the full model. For PFNA, energy intake, dairy, eggs, popcorn, income to poverty ratio, parity and alcohol were removed compared to the full model.

Examination of the percent difference in PFAS by quartile of fiber showed a roughly linear relationship for PFOS and PFNA (Figure 1). For PFOA, the highest quartile had a smaller percent difference than would be consistent with a linear relationship. This may represent some type of saturation of binding at higher fiber levels, though the quadratic model did not provide a substantially better fit to the data.

**Figure 1.**
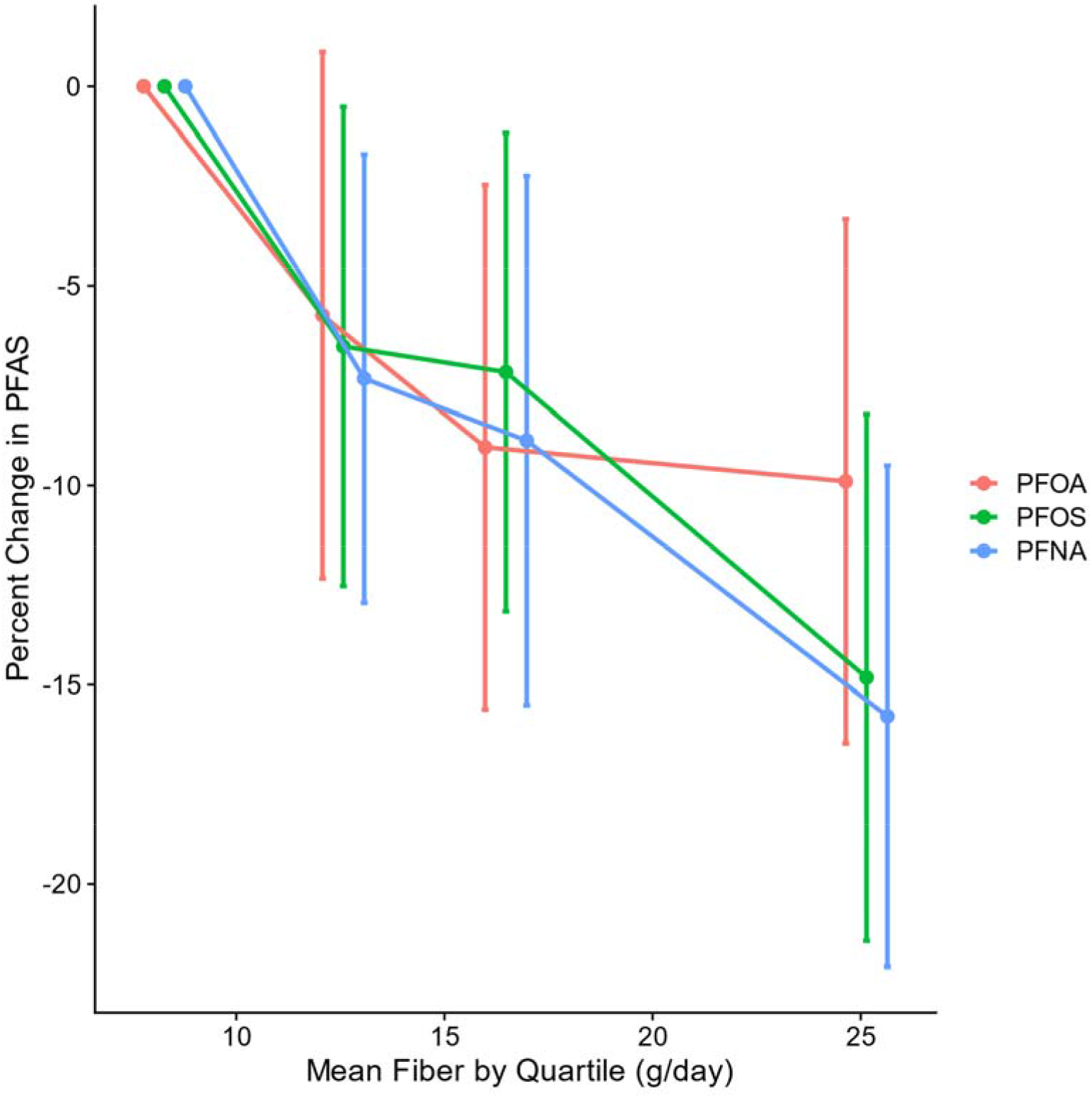
Percent change in PFAS by quartile of energy-adjusted dietary fiber.^†^ ^†^ Energy adjusted mean fiber values by quartile were 8.27, 12.6, 16.5, and 25.1 g/day.

The effect modification analysis showed interaction for age (PFOA and PFNA), BMI (all three PFAS), energy intake (PFOS), smoking category (PFOS and PFNA), alcohol category (PFOS), and NHANES wave (PFOA) (Tables S6 and S7a-c). Fiber associations decreased with age (PFOA and PFNA). Fiber associations decreased with BMI for PFOA; for PFOS and PFNA the trend was in the opposite direction and for PFNA, was of smaller magnitude. The association with fiber increased with energy intake (PFOS). Heavy smokers [≥1 pack per day average] showed no decrease in PFOS or PFNA with higher fiber intake. The fiber associations in later waves were less inverse (PFOA and PFNA). The inverse association with fiber was largest for never, light and moderate drinkers (PFOS). Although differences in the PFAS-fiber association by sex did not meet our criterion for statistical significance, the sex-stratified results suggested that the association for PFOA and PFOS was more inverse among females (Table S8).

For PFOA the analysis of type of food contributing fiber did not support important differences in association, based on the AIC’s of the models (Table S9). The PFOS-fiber association was most inverse for fiber from the fruit food group. The PFNA model also supported differences by fiber source; in this case fruit and grain-based mixed dishes had the most inverse relations. When we examined associations with g/d of food from fiber containing foods, for PFOA, the model comparison supported a difference among food sources, and the food category highest in fiber (plant-based protein foods, which includes beans) had the most inverse relation (Table S10). Note that for PFOA, the g/d fiber models fit better than the g/d food models. For PFNA, the g/d food results were like those for PFOA. For PFOS, the g/d food models did not support a difference by food groups, and the total g/d food model fit better than the total g/d fiber model. Multiple imputation showed little difference from the complete case analysis (Table S11).

The measurement error-adjusted estimates for total fiber, as expected, were more inverse than those shown in Table 3 (supplemental information, section 1). The adjusted estimated were more inverse by 75% for PFOA, 82% for PFOS, and 53% for PFNA. When intake of crude fiber was modeled instead of energy-adjusted fiber, the results for crude fiber were similar (Table S12).

## Discussion

For all three PFAS examined, higher intake of dietary fiber was associated with lower PFAS concentrations in serum. Although the associations were of modest size (less than 10% decrease in PFAS with IQR increase in dietary fiber), they may be important in the context of outcome-PFAS associations that themselves are of modest size, for which unmeasured confounding may have an important influence on the results.

In 13 previous reports with information about serum PFAS concentrations in relation to dietary factors, the focus was primarily on foods that might increase blood levels (Arrebola et al., 2018; Averina et al., 2018; Brantsaeter et al., 2013; Christensen et al., 2017; Eriksen et al., 2011; Halldorsson et al., 2008; Jain, 2014; Liu et al., 2017; Park et al., 2019; Skuladottir et al., 2015; Sun et al., 2018; Wu et al., 2015; Zhou et al., 2019). Dietary fiber was not examined in these studies, although associations of PFAS with intake of fiber-rich foods such as fruits, vegetables, beans, and cereal were considered by many. In a more recent study, Lin et al. examined both dietary patterns and energy-adjusted fiber intake directly and found high fiber diet patterns and fiber were associated with lower levels of several PFAS (Lin et al., 2020). Relevant to the interpretation of the data on foods is that energy intake is an expected confounder of the PFAS-fiber relationship and without adjustment an inverse association would be harder to detect. Adjustment for energy intake was done in three the of previous studies (Brantsaeter et al., 2013; Lin et al., 2020; Skuladottir et al., 2015). Despite the lack of energy intake adjustment in most of the previous studies that examined associations of PFAS with fiber-rich foods, the results were generally consistent with an inverse relation. In a study based on the Danish National Birth Cohort (Halldorsson et al., 2008), PFOA and PFOS were inversely related to intake of vegetables, and PFOS was inversely related to intake of fruits. In a study of 2003-2008 NHANES subjects (Jain, 2014), PFOA was inversely related to intake of beans, despite using only one diet record per subject, which would decrease the ability to detect a relation. In a study done in Singapore (Liu et al., 2017), PFOA and PFOS were inversely related to grains, and PFOS was inversely related to soy products. In another study done in Denmark (Skuladottir et al., 2015), PFOA and PFOS were inversely related to vegetable intake. In the Nurses’ Health Study (Sun et al., 2018), an index of healthy eating, which was likely correlated with fiber intake, was related to lower PFOS. In a study done in California (Wu et al., 2015), PFOS concentration was inversely related to intake of crackers (excluding graham crackers). Popcorn was the only high-fiber food with a positive relation (Park et al., 2019; Sun et al., 2018), probably because the packaging for some popcorn products was a source of exposure (Sinclair et al., 2007).

The analysis of the PFAS-fiber association according to fiber source indicated that fiber’s inverse association with PFOS and PFNA varied across food groups (Table S10). These analyses suggested that fruit fiber was especially important. For PFOA, no such specificity was indicated. A variety of fiber sources may be important. Although the g/d fiber models fit the data better than the models of g/d food from fiber-rich food groups for PFOA and PFNA, for PFOS this was not the case. Perhaps the g/d food model fit better for PFOS because it did a better job of capturing PFOS exposure from food packaging material (Susmann et al., 2019). The broad soluble/insoluble fiber dichotomy once used has been declared obsolete by the National Academy of Sciences, and for that reason even this breakdown has not been implemented in the USDA database on which the NHANES nutrient information was based (Slavin, 2003).

The effect modification analysis identified no factor that similarly altered the PFAS-fiber association across all three PFAS. Age, BMI, and smoking, however, did similarly alter the association for at least two PFAS. The obese tend to underreport energy intake and distort reporting of dietary composition (Heitmann et al., 1995; Lissner et al., 2007). The net effect could be over-reporting of fiber intake among the obese. In other words, the true fiber intake among the obese could be less than reported, giving the appearance of a diminished fiber effect. Age-related differences in the accuracy of reported diet could similarly distort our results (Krok-Schoen et al., 2019). Given the limited evidence pointing to specific food sources of fiber as being important, co-variation in fiber type consumed with heavy smoking, age or BMI appears to be less well supported as an explanation for the effect modification but is still possible.

As noted earlier, dietary fiber decreases absorption of several constituents of food such as cholesterol, fat, and iron (Grube et al., 2013; Hallberg and Hulthén, 2000; Jesch and Carr, 2017). Dietary fiber decreases absorption by adsorbing substances and trapping them in a viscous gel matrix (Naumann et al., 2019). The association between dietary fiber and PFAS may be due to a similar underlying mechanism.

Although PFAS enters the gut by consuming food and drinking water that contain it, PFAS also enters the gut in bile that is excreted as part of enterohepatic circulation. This has been shown through measurement of PFOA and PFOS in bile at concentrations similar to those found in serum (Harada et al., 2007), and identification of many of the transporters that facilitate their enterohepatic circulation (Zhao et al., 2017, 2015). PFAS entering the gut through bile would be subject to the same effects of dietary fiber, i.e. decreased reabsorption, as PFAS from food. An analogous effect occurs with ingestion of cholestyramine, a bile acid sequestrant that also binds to selected drugs and nutrients (Davies, 1998; Pieroni and Fisher, 1981; West and Lloyd, 1975), which has been shown to increase excretion in human feces of all three PFAS we studied (Genuis et al., 2010). In that case study, excretion of PFOS and PFNA were increased to a greater degree than for PFOA (Genuis et al., 2010). Because we examined a small number of PFAS compounds, our ability to make inferences about the relationship of structure to fiber-enhanced gastrointestinal excretion was limited. Nonetheless, our results suggest that for fiber-enhanced excretion, alkyl chain length may be more important than the attached functional group.

Fiber intake may be an important unmeasured confounder affecting epidemiologic associations between serum cholesterol and serum PFAS, as mentioned previously. Fiber decreases serum cholesterol (Brown et al., 1999), and appears to lower serum PFAS. Some epidemiologic associations between serum cholesterol and serum PFAS could be attenuated by adjustment for fiber intake. Binding of fiber to PFAS that is in the gut due to enterohepatic circulation could account for confounding in some studies where PFAS exposure is primarily via drinking water or occupation (Steenland et al., 2009; Costa et al., 2009). Fiber could also be a confounder in epidemiologic associations between immunogenicity and PFAS. Fiber intake in the form of prebiotic supplements has been shown in randomized clinical trials to increase antibody titers following influenza vaccination (Yeh et al., 2018). If fiber increases antibody response to vaccines, and also decreases PFAS, this could account for some of the inverse association of antibody response with PFAS in epidemiologic data, but not the immune toxicity in experimental data (Liu et al., 2020; McDonough et al., 2020). The method of quantifying dietary fiber used for the USDA database underlying NHANES 2005-2016 did not include prebiotic dietary fiber as a separate category from total dietary fiber (Phillips et al., 2019). Total dietary fiber, however, did capture dietary constituents that -- like prebiotics -- increase the production of short chain fatty acids, which have favorable effects on systemic antibody response (Carlson et al., 2018; Kim et al., 2016). Assessing the degree to which these associations are confounded by fiber will not be easy given the difficulty of measuring diet well. For example, if we had examined the cholesterol-PFAS association after adjusting for fiber intake and other potential confounders in the NHANES data, measurement error in the estimated usual fiber intake would lead to an overestimate of the association due to residual confounding. Specialized methods that account for the measurement error would be needed to estimate an unconfounded association.

Because our assessment of diet was based on two 24-hour diet recalls and because of the day-to-day variation in diet, we had an imprecise measure of usual intake for each subject. Imprecision in the fiber measure means that the associations with PFAS we observed were attenuated. We estimated the effect of imprecision on the associations by taking the correlation between the two energy-adjusted fiber intakes for each subject into account; this suggested that the estimates for the fiber “effect” would have been more than 70% larger for PFOA and PFOS in the absence of imprecision (supplemental information, section 1). The imprecision also applied to the dietary confounders, meaning that the fiber estimate might have been influenced by residual confounding. For example, meat was directly associated with PFOA (Table S1) and was inversely associated with fiber (Table 2). Failure to measure meat intake precisely means that the PFAS-fiber association would appear to be more inverse than in fact (Mehio-Sibai et al., 2005). Although meat was not a statistically significant determinant of serum PFOA, the possibility exists that the estimates in Table 3 were larger on an absolute scale than in fact. For the other dietary confounders, based on the sign of their relation with fiber and with PFOA, the residual confounding would have caused an estimate for fiber that was biased towards the null. What the overall effect of the residual confounding was remains uncertain. Another level of uncertainty arose from the inaccuracy in 24-hour diet recalls (Karvetti and Knuts, 1985), which would have added to the imprecision and could have biased the results. Despite the difficulty incurred by use of a dietary measure, we were able to detect the modest associations, possibly because of the large sample size, and the precision of the PFAS assays. Nonetheless, confirmation of the PFAS-fiber association in large studies with a comprehensive diet assessment and data analysis would increase confidence.

## Conclusion

These results suggest that dietary fiber increases the gastrointestinal excretion of PFOA, PFOS, and PFNA in humans. Although the size of the associations was modest (less than 10% difference in PFAS with IQR difference in dietary fiber), the findings may be important in the context of studies of health outcomes in relation to PFAS if those health outcomes are also related to dietary fiber intake and the outcome-PFAS association is also modest.

## Data Availability

Data is available upon request.

## CRediT authorship contribution statement

Michael W. Dzierlenga: Methodology, Software, Investigation, Writing – original draft & editing. Debra R. Keast: Methodology, Writing – review and editing.

Matthew P. Longnecker: Conceptualization, Funding acquisition, Methodology, Writing - review & editing.

