## Supplemental Information for "The concentration of several perfluoroalkyl acids in serum appears to be reduced by dietary fiber"

Michael W. Dzierlenga^a,*^, Debra R. Keast^b^, Matthew P. Longnecker^a^

^a^ Ramboll, Raleigh, NC, USA

^b^ Food & Nutrition Database Research, Inc., Bangor, PA, USA

### 1. Estimation of the PFAS-fiber association in the absence of measurement error

As noted in the main report, use of two 24-hour dietary recalls provided an imprecise measure of usual fiber intake, due to the day-to-day variation in diet. Because we had two 24-hour dietary recalls for each subject, we were able to estimate what the PFAS-fiber association would have been in the absence of this imprecision.

We could de-attenuate the initial estimate of *𝛽_fiber_* based on the fiber intake in one recall but not if we had used the average fiber based on two recalls. So, we fit the models of PFAS using the first 24-hour recall data. We then estimated *𝛽_fiber-DA_* (the de-attenuated estimate) using *𝛽_fiber_* as follows (Keogh et al. 2020):

*𝛽_fiber-DA_* = *𝛽_fiber_*/*r*

Where *r* is the Pearson correlation coefficient between the energy-adjusted fiber values of the two 24-hour recalls. Here we are treating the two 24-hour recalls as a replicates study. Because we have a large sample size for the replicates study (n=6,482), we assumed no additional variance in the estimate of *𝛽_fiber-DA_* due to the de-attenuation (Fibrinogen Studies Collaboration, 2009), and assumed *𝛽_fiber-DA_* had the same *t* as *𝛽_fiber_*. We then repeated this procedure using the second 24-hour recall data, and calculated the inverse variance mean of the two results.

*𝛽_fiber_* (and 95% CI) and corresponding % difference in PFAS (and 95% CI)

obtained using one 24-hour diet recall*

|  | *𝛽_fiber_* | %Δ in PFAS |
| --- | --- | --- |
| PFOA | -0.0305 (-0.0554, -0.0057) | -3.01 (-5.39, -0.56) |
| PFOS | -0.0603 (-0.0891, -0.0315) | -5.85 (-8.53, -3.10) |
| PFNA | -0.0703 (-0.0999, -0.0407) | -6.79 (-9.51, -3.99) |

*The model of PFAS used was otherwise the same as the “final” model in the main report (n=6,482); weighted mean results from the first and second recalls are shown.

Pearson correlation coefficients between energy-adjusted dietary fiber from the two diet recalls (n=6,482)

| No transformation in energy-adjusted fiber | Box Cox transformation of energy-adjusted fiber |
| --- | --- |
| 0.467 | 0.465 |

De-attenuated estimates of % difference in PFAS (and 95% CI),

with comparison to values based on the average of two 24-hour recalls

|  | %Δ in PFAS*_fiber-DA_* | %Δ in PFAS*_fiber-2 24-h_** | % increase |
| --- | --- | --- | --- |
| PFOA | -6.35 (-11.28, -1.16) | -3.64 | 75 |
| PFOS | -12.16 (-17.40, -6.60) | -6.69 | 82 |
| PFNA | -12.80 (-18.96, -6.18) | -8.36 | 53 |

* These are the same results shown for the final model in Table 3.

The IQD in energy-adjusted fiber intake for the de-attenuated estimates is 8.4 g/d of energy adjusted fiber, as compared with the values of 8.3 for the IQD based on the average of two 24-hour diet recalls. The IQD in in energy-adjusted fiber intake was calculated with the NRC method (Shaw et al., 2020), using Box Cox transformed values and the distribution of energy-adjusted fiber in the first 24-hour recall.

Comments:

The two-day estimate for PFNA (-8.36%) was less attenuated than for the other two PFAS, despite being subject to the same error-generating process. The weights that account for the day of the week were used in the main analysis but not in the error-correction calculations. If the weights had been used in the error correction calculations, the de-attenuated coefficients would have been more negative, especially for PFNA.

Keogh RH, Shaw PA, Gustafson P, Carroll RJ, Deffner V, Dodd KW, Küchenhoff H, Tooze JA, Wallace MP, Kipnis V, Freedman LS. STRATOS guidance document on measurement error and misclassification of variables in observational epidemiology: Part 1-Basic theory and simple methods of adjustment.

Stat Med. 2020 Jul 20;39(16):2197-2231.

Shaw PA, Gustafson P, Carroll RJ, Deffner V, Dodd KW, Keogh RH, Kipnis V, Tooze JA, Wallace MP, Küchenhoff H, Freedman LS. STRATOS guidance document on measurement error and misclassification of variables in observational epidemiology: Part 2-More complex methods of adjustment and advanced topics. Stat Med. 2020 Jul 20;39(16):2232-2263.

2. Supplemental Tables & Figures

Table S1. Classification of individual food recall items into categories for analysis.

| Food Category | USDA Food Code^a^ | Description |
| --- | --- | --- |
| Milk & Milk Products | 111* | Milk |
|  | 112* | Evaporated & Condensed Milk |
|  | 114* | Yogurt |
|  | 115* | Milk Treats (e.g. Milk Shakes, Hot Chocolate, etc.) |
|  | 118* | Dried Milk, Not Reconstituted |
|  | 12[13]* | Cream; Sour Cream |
|  | 13[1234]* | Ice Cream; Pudding; Pudding, Baby-food; White Sauces |
|  | 14* | Cheese |
|  | 81101* | Butter |
|  | 81105* | Butter Blends |
|  | 81106* | Butter Replacement |
|  | 81204* | Clarified Butter |
| Meat & Meat Products | 20* | Meat, Not Further Specified |
|  | 21* | Beef |
|  | 22* | Pork |
|  | 23* | Lamb, Veal & Game Meat |
|  | 24* | Poultry |
|  | 25* | Organ meats, Sausage & Lunchmeat |
|  | 27[1234]1* | Dishes Containing Beef |
|  | 27[1234]2* | Dishes Containing Pork |
|  | 27[1234]3* | Dishes Containing Lamb, Veal & Game Meat |
|  | 27[1234]4* | Dishes Containing Poultry |
|  | 27[1234]6* | Dishes Containing Organ Meats, Sausage & Lunchmeat |
|  | 275[12467]* | Sandwiches Containing Meat (Not Fish/Seafood) |
|  | 276* | Baby-Foods Containing Meat |
|  | 281[1346]* | Frozen Meals Containing Meat |
|  | 283[12346]* | Soups Containing Meat |
| Eggs | 3[12]* | Eggs and Dishes Containing Eggs |
| Popcorn | 54403* | Popcorn and Caramel Corn |
|  | 54319020 | Popcorn Cake |

^a^ Where * serves as a ‘wildcard’ character and square brackets serve as a logical “or”. For example, 111* represents any food code beginning with 111, which are in that case several different types of milk. 2[012]* represents the food categories represented by 20*, 22* and 21*. Categories which are combined using square brackets are separated by a semi-colon in the description field.

Figure S1. Directed acyclic graph of the effect of dietary fiber on PFAS concentration in serum. BMI, body mass index; IPR, income to poverty ratio; Wave refers to NHANES sampling period; EtOH, alcohol consumption; EI, energy intake. Diet composition here is used as a broad term that encompasses not only elements of the diet that contain PFAS, but also the qualitative nature of diet; e.g., diets high in PFAS-containing foods may include less fiber. Alcohol consumption would also affect energy intake, but to simplify the figure, no arrow is shown.

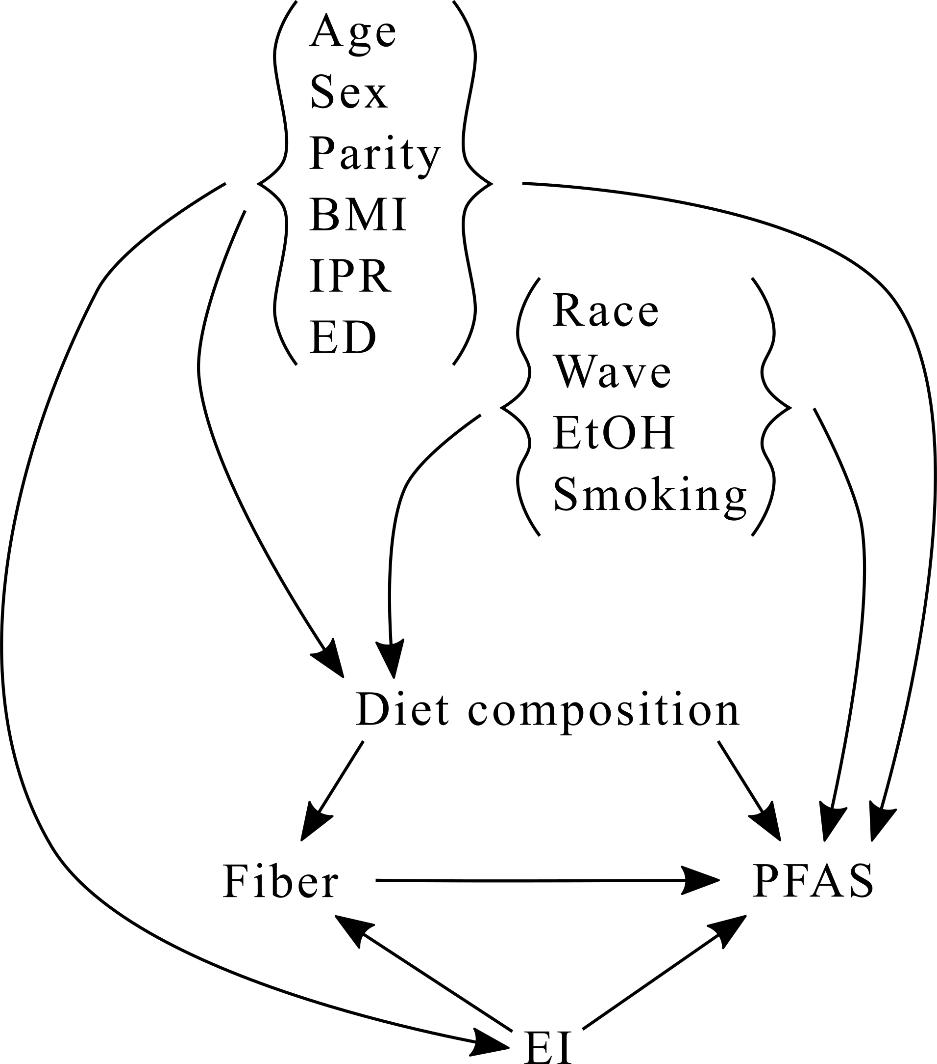

Table S2. Mean Dietary Fiber (g/d) Intake Contributed According to Food Categories used in the Analysis^a^

|  |  |  | 2-Day Average Fiber (g) Intake | | | |
| --- | --- | --- | --- | --- | --- | --- |
| Food Category Description | | | Mean | ± | SE | Pct (%) |
| All Food Groups | | | 17.13 | ± | 0.20 | 100.00 |
| Fruit | |  | 1.81 | ± | 0.05 | 10.59 |
| Vegetables | | | 3.07 | ± | 0.06 | 17.93 |
| Plant-based Protein Foods | | | 1.61 | ± | 0.08 | 9.42 |
| Grain-based Mixed Dishes^b^ | | | 0.87 | ± | 0.04 | 5.10 |
| Savory snacks, crackers, cereal/nutrition bars, and baked goods | | | 1.66 | ± | 0.04 | 9.66 |
| Savory snacks and crackers | | | 0.86 | ± | 0.03 | 5.03 |
|  | Savory snacks | | 0.69 | ± | 0.03 | 4.04 |
|  |  | Potato chips | 0.16 | ± | 0.01 | 0.94 |
|  |  | Tortilla, corn, other chips | 0.25 | ± | 0.02 | 1.43 |
|  |  | Popcorn | 0.22 | ± | 0.02 | 1.29 |
|  |  | Pretzels/snack mix | 0.07 | ± | 0.01 | 0.38 |
|  | Crackers | | 0.17 | ± | 0.01 | 0.99 |
| Cereal/nutrition bars and baked goods | | | 0.79 | ± | 0.03 | 4.63 |
|  | Cereal/nutrition bars | | 0.21 | ± | 0.02 | 1.20 |
|  |  | Cereal bars | 0.14 | ± | 0.02 | 0.80 |
|  |  | Nutrition bars | 0.07 | ± | 0.01 | 0.40 |
|  | Baked goods | | 0.59 | ± | 0.02 | 3.43 |
|  |  | Cakes and pies | 0.22 | ± | 0.01 | 1.28 |
|  |  | Cookies and brownies | 0.23 | ± | 0.01 | 1.36 |
|  |  | Doughnuts, sweet rolls, pastries | 0.14 | ± | 0.01 | 0.80 |
| Other fiber sources | | | 8.10 | ± | 0.14 | 47.28 |

^a^ NHANES, 2005-2016, 2-day dietary sample, ages 20+ yr, with serum perfluoralkyl substances (PFAS) measurement and included in the regression analyses. Sample-weighted mean and standard error were estimated using SUDAAN.

^b^ Includes grain foods, grain-based mixed dishes, pizza, and sandwiches. Does not include grain-based snacks & sweets.

Figure S2. Flowchart showing the total population in the combined NHANES waves and the exclusions applied to generate the population used in our analysis.

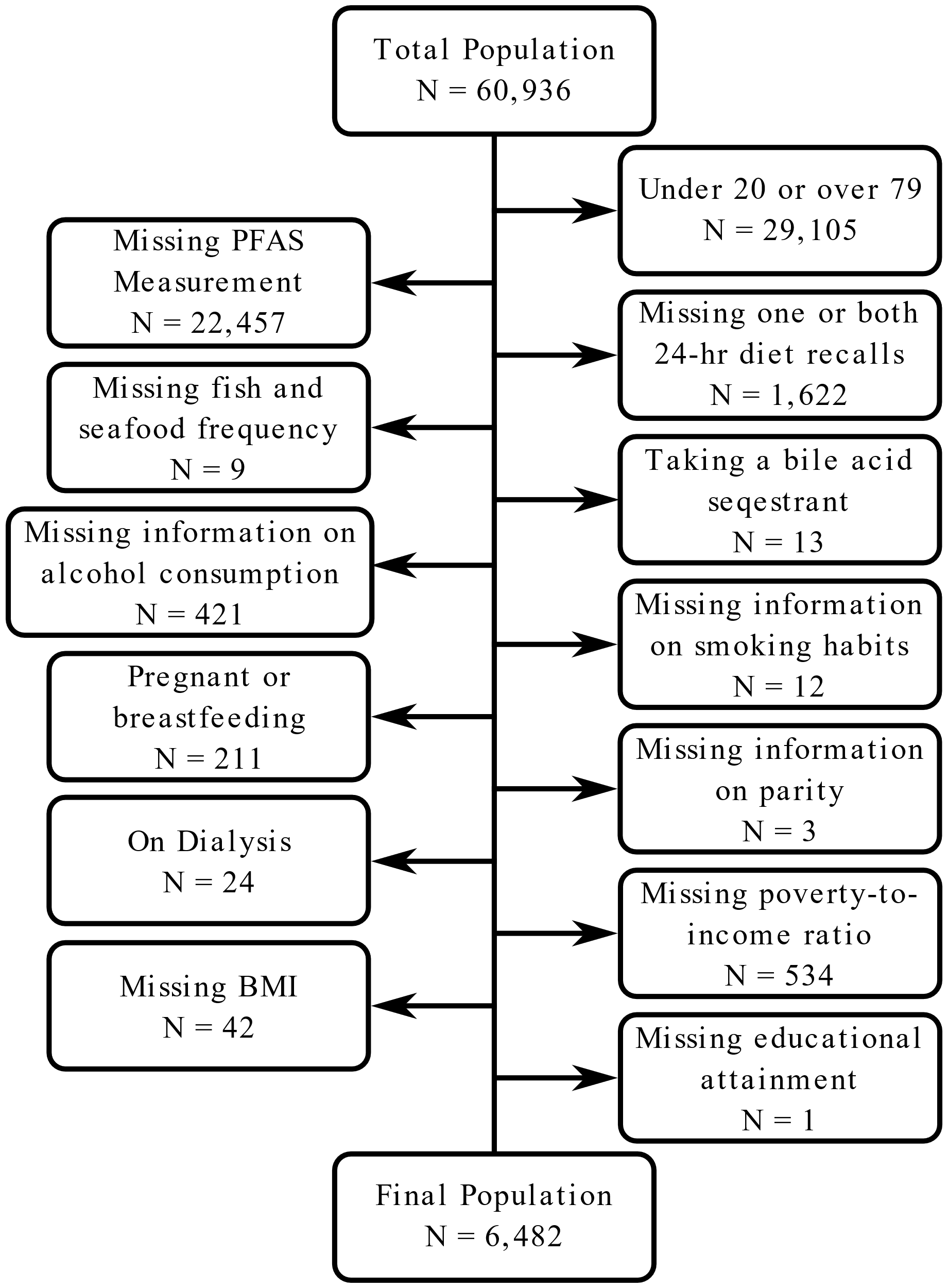

Table S3. Set of β coefficients for regression with PFOA. (Only variables in final model)

| Variable | β (95% CI) |
| --- | --- |
| Intercept | 1.21e+00 (1.01e+00, 1.40e+00) |
| Dietary Fiber (inter-quartile shift) | -3.71e-02 (-6.35e-02, -1.07e-02) |
| Sex (female as referent) | 1.68e-01 (1.05e-01, 2.30e-01) |
| Age | 7.26e-03 (5.90e-03, 8.63e-03) |
| Race (Mexican American) | -2.41e-01 (-3.22e-01, -1.60e-01) |
| Race (Other Hispanic) | -1.50e-01 (-2.35e-01, -6.51e-02) |
| Race (Non-Hispanic Black) | -1.54e-01 (-2.16e-01, -9.25e-02) |
| Race (Other Race) | -1.10e-01 (-1.80e-01, -4.04e-02) |
| Wave | -1.49e-01 (-2.29e-01, -6.88e-02) |
| Wave^2^ | -1.05e-02 (-2.47e-02, 3.68e-03) |
| BMI (kg/m^2^) | -5.33e-03 (-8.48e-03, -2.17e-03) |
| Education (Some high school) | 6.86e-02 (-2.80e-02, 1.65e-01) |
| Education (High school diploma) | 1.04e-01 (1.79e-02, 1.91e-01) |
| Education (Some College or AA degree) | 9.13e-02 (-4.49e-03, 1.87e-01) |
| Education (College degree) | 7.08e-02 (-1.82e-02, 1.60e-01) |
| Parity (1) | -9.76e-02 (-1.86e-01, -9.46e-03) |
| Parity (2 or more) | -1.48e-01 (-2.12e-01, -8.49e-02) |
| Alcohol (former) | -8.69e-03 (-9.47e-02, 7.73e-02) |
| Alcohol (light drinker) | 1.57e-02 (-5.74e-02, 8.89e-02) |
| Alcohol (drinker) | 6.52e-02 (-9.60e-03, 1.40e-01) |
| Alcohol (heavy drinker) | 6.30e-02 (-2.35e-02, 1.49e-01) |
| Fish & Shellfish Consumption (occurrences last 30 d) | 5.08e-03 (2.52e-03, 7.63e-03) |
| Meat Consumption (g/day) | 1.10e-04 (-7.27e-05, 2.93e-04) |
| Popcorn Consumption (g/day) | 2.37e-03 (5.78e-04, 4.16e-03) |

Table S4. Set of β coefficients for regression with PFOS. (Only variables in final model)

| Variable | β (95% CI) |
| --- | --- |
| Intercept | 2.38e+00 (2.21e+00, 2.55e+00) |
| Dietary Fiber (inter-quartile shift) | -6.93e-02 (-1.01e-01, -3.80e-02) |
| Sex (female as referent) | 4.59e-01 (4.19e-01, 4.99e-01) |
| Age | 1.31e-02 (1.15e-02, 1.47e-02) |
| Race (Mexican American) | -2.12e-01 (-2.86e-01, -1.39e-01) |
| Race (Other Hispanic) | -2.27e-01 (-3.33e-01, -1.22e-01) |
| Race (Non-Hispanic Black) | 1.53e-01 (8.09e-02, 2.25e-01) |
| Race (Other Race) | -2.25e-02 (-1.14e-01, 6.86e-02) |
| Wave | -2.70e-01 (-2.86e-01, -2.54e-01) |
| BMI (kg/m^2^) | -8.68e-03 (-1.18e-02, -5.57e-03) |
| Education (Some high school) | -4.43e-03 (-1.10e-01, 1.01e-01) |
| Education (High school diploma) | 5.13e-02 (-4.56e-02, 1.48e-01) |
| Education (Some College or AA degree) | 4.90e-02 (-4.66e-02, 1.45e-01) |
| Education (College degree) | -2.81e-03 (-9.78e-02, 9.22e-02) |
| Smoking (former) | -7.20e-02 (-1.19e-01, -2.48e-02) |
| Smoking (smoker) | -1.93e-01 (-2.60e-01, -1.27e-01) |
| Smoking (heavy smoker) | -1.73e-01 (-2.49e-01, -9.78e-02) |
| Fish & Shellfish Consumption (occurrences last 30 d) | 8.92e-03 (5.84e-03, 1.20e-02) |

Table S5. Set of β coefficients for regression with PFNA. (Only variables in final model)

| Variable | β (95% CI) |
| --- | --- |
| Intercept | -1.48e-02 (-2.21e-01, 1.91e-01) |
| Dietary Fiber (inter-quartile shift) | -1.31e-01 (-1.93e-01, -6.86e-02) |
| Dietary Fiber^2^ (inter-quartile shift) | 4.33e-02 (1.31e-03, 8.53e-02) |
| Sex (female as referent) | 1.46e-01 (1.13e-01, 1.80e-01) |
| Age | 7.44e-03 (5.60e-03, 9.28e-03) |
| Race (Mexican American) | -6.57e-02 (-1.54e-01, 2.31e-02) |
| Race (Other Hispanic) | 2.76e-02 (-7.09e-02, 1.26e-01) |
| Race (Non-Hispanic Black) | 1.19e-01 (5.18e-02, 1.87e-01) |
| Race (Other Race) | 7.33e-02 (-1.16e-02, 1.58e-01) |
| Wave | 8.93e-02 (-2.87e-02, 2.07e-01) |
| Wave^2^ | -4.76e-02 (-6.79e-02, -2.73e-02) |
| BMI (kg/m^2^) | -5.95e-03 (-9.43e-03, -2.48e-03) |
| Education (Some high school) | -2.85e-02 (-1.08e-01, 5.13e-02) |
| Education (High school diploma) | 2.74e-02 (-4.90e-02, 1.04e-01) |
| Education (Some College or AA degree) | -2.62e-02 (-1.12e-01, 6.00e-02) |
| Education (College degree) | 3.72e-03 (-7.95e-02, 8.70e-02) |
| Smoking (former) | -2.41e-02 (-8.08e-02, 3.25e-02) |
| Smoking (smoker) | -6.94e-02 (-1.22e-01, -1.72e-02) |
| Smoking (heavy smoker) | -7.71e-02 (-1.66e-01, 1.20e-02) |
| Fish & Shellfish Consumption (occurrences last 30 d) | 1.44e-02 (1.07e-02, 1.81e-02) |
| Meat Consumption (g/day) | 1.89e-04 (5.68e-05, 3.22e-04) |

Table S6. Percent change in PFAS by level of interaction variable (continuous variables).^*^

| PFAS | Interaction Variable | 25^th^ Percentile | 50^th^ Percentile | 75^th^ Percentile |
| --- | --- | --- | --- | --- |
| PFOA | Age | -6.56 (-3.30, -9.72) | -4.18 (-1.60, -6.69) | -2.11 (0.87, -5.00) |
|  | BMI | -5.44 (-8.27, -2.53) | -3.97 (-6.42, -1.46) | -2.04 (-4.81, 0.82) |
| PFOS | BMI | -5.29 (-2.12, -8.35) | -6.85 (-3.93, -9.67) | -8.90 (-5.64, -12.04) |
|  | Energy | -8.46 (-5.06, -11.75) | -6.91 (-3.94, -9.79) | -4.87 (-1.80, -7.85) |
| PFNA | Age | -10.01 (-7.01, -12.91) | -7.98 (-5.34, -10.55) | -6.23 (-2.67, -9.66) |
|  | BMI | -7.92 (-4.65, -11.07) | -8.14 (-5.45, -10.74) | -8.42 (-5.61, -11.15) |

^*^The table entries show the percent change in PFAS per interquartile distance increment in fiber intake (and 95% confidence interval), according to the percentile of the interaction variable. A quadratic model of PFNA was used. Interaction was examined using the full model with all covariates.

Table S7a-c. Percent change in PFAS per interquartile distance increment in fiber intake, with effect modification by category of modifying variable. Interaction was examined using the full model with all covariates.

a. Smoking

| PFAS | Nonsmoker | Former Smoker | Smoker | Heavy Smoker |
| --- | --- | --- | --- | --- |
| PFOS | -8.78 (-12.34, -5.08) | -5.23 (-9.99, -0.22) | -2.05 (-8.67, 5.06) | 10.29 (-2.80, 25.13) |
| PFNA | -10.28 (-7.53, -12.96) | -8.55 (-5.74, -11.28) | -6.47 (-3.59, -9.26) | 14.07 (17.58, 10.67) |

b. Wave

|  | | %∆ per IQD increment in fiber |
| --- | --- | --- |
| Wave | PFOA | PFNA |
| 2005-2006 | -8.04 (-14.42, -1.19) | -11.12 (-8.38, -13.77) |
| 2007-2008 | -3.86 (-9.55, 2.19) | -10.90 (-8.17, -13.56) |
| 2009-2010 | -7.40 (-12.06, -2.49) | -11.92 (-9.22, -14.55) |
| 2011-2012 | 0.12 (-5.58, 6.16) | -6.16 (-3.28, -8.96) |
| 2013-2014 | -5.44 (-12.19, 1.83) | -2.74 (0.25, -5.64) |
| 2015-2016 | 2.56 (-2.98, 8.43) | -3.91 (-0.95, -6.77) |

c. Alcohol

| PFAS | Category^*^ | %∆ per IQD increment in fiber |
| --- | --- | --- |
| PFOS | Never | -9.43 (-16.42, -1.84) |
|  | Former | 0.41 (-6.15, 7.43) |
|  | Light | -8.73 (-13.78, -3.38) |
|  | Drinker | -8.03 (-11.98, -3.90) |
|  | Heavy | -1.67 (-7.88, 4.97) |

* See Table 1 for exact category definitions

Table S8. Sex-stratified percent change in PFAS level per IQR increase in dietary fiber intake. ^†^

|  |  | Percent Change (95% CI) | | |
| --- | --- | --- | --- | --- |
| Sex | PFAS (ng/mL) | Crude | Full | Final |
| Male | PFOA | -4.91 (-8.17, -1.54) | -2.77 (-5.26, -0.22) | -2.19 (-4.66, 0.35) |
|  | PFOS | -4.45 (-8.64, -0.08) | -5.42 (-8.96, -1.74) | -5.39 (-8.79, -1.86) |
|  | PFNA | -6.78 (-10.73, -2.66) | -8.59 (-11.75, -5.31) | -8.63 (-12.50, -4.59) |
| Female | PFOA | -2.62 (-6.94, 1.90) | -5.16 (-9.51, -0.59) | -5.28 (-9.46, -0.90) |
|  | PFOS | -5.07 (-9.95, 0.08) | -8.03 (-12.32, -3.52) | -7.91 (-12.21, -3.40) |
|  | PFNA | -3.63 (-8.34, 1.31) | -7.15 (-11.77, -2.28) | -7.92 (-12.41, -3.19) |

^†^ The full model was adjusted for energy intake, consumption of meat and meat products, milk and dairy products, eggs, popcorn, fish and shellfish, age, BMI, income to poverty ratio, education, parity (in women), sex, NHANES wave, smoking, alcohol, and race/ethnicity. The final model was based on the set of adjustment variables which gave a % change in PFAS that was < 5% different from that in the full model. For PFOA, energy intake, dairy, eggs, income to poverty ratio and smoking were removed. For PFOS, energy intake, meat, dairy, eggs, popcorn, income-to-poverty ratio, parity, wave^2^ and alcohol were removed compared to the full model. For PFNA, energy intake, dairy, eggs, popcorn, income-to-poverty ratio, parity and alcohol were removed compared to the full model.

Table S9. Percent change in PFAS per g/d fiber, for total fiber, and for fiber from specific food categories^a^

|  |  |  | Percent change in PFAS with g/day increase in fiber | | | | | | |
| --- | --- | --- | --- | --- | --- | --- | --- | --- | --- |
|  |  | AIC | Total | Fruit | Vegetable | Plant-based Protein Foods | Grain-based Mixed Dishes^b^ | Grain-based Snacks & Sweets | Other Food Sources |
| PFOA | Baseline | 14096.09 | -0.45  (-0.77, -0.13) | NA | NA | NA | NA | NA | NA |
|  | Fiber Sources | 14096.31 | NA | -0.87  (-1.63, -0.11) | -0.63  (-1.24, -0.01) | -0.54  (-1.12, 0.03) | -0.19  (-0.62, 0.23) | -0.35  (-1.55, 0.85) | -0.28  (-0.88, 0.31) |
| PFOS | Baseline | 15653.99 | -0.67  (-1.03, -0.31) | NA | NA | NA | NA | NA | NA |
|  | Fiber Sources | 15644.88 | NA | -1.55  (-2.42, -0.67) | -0.59  (-1.49, 0.32) | -0.49  (-1.14, 0.17) | -0.72  (-1.21, -0.24) | -1.04  (-2.13, 0.07) | -0.81  (-1.34, -0.26) |
| PFNA | Baseline | 14491.88 | -1.51  (-2.23, -0.78) | NA | NA | NA | NA | NA | NA |
|  | Fiber Sources | 14484.07 | NA | -1.91  (-2.96, -0.86) | -0.52  (-1.49, 0.47) | -0.52  (-1.58, 0.56) | -1.90  (-2.70, -1.09) | -0.18  (-2.16, 1.84) | -1.45  (-2.14, -0.76) |

^a^ Adjustment factors are the same as for the final model in Table 3, except that total energy intake was also included as a covariate (n=6,482).

^b^ Includes grain foods, grain-based mixed dishes, pizza, and sandwiches. Does not include grain-based snacks & sweets.

Table S10. Percent change in PFAS per g/d of food from fiber containing foods^a^

|  |  |  | Percent change in PFAS with g/day increase in food intake | | | | | | |
| --- | --- | --- | --- | --- | --- | --- | --- | --- | --- |
|  |  | AIC | Total | Fruit | Vegetable | Plant-based Protein Foods | Grain-based Mixed Dishes^b^ | Grain-based Snacks & Sweets | Other Food Sources |
| PFOA | Baseline | 14109.63 | -0.00  (-0.00, 0.00) | NA | NA | NA | NA | NA | NA |
|  | Fiber Sources | 14101.12 | NA | -0.01  (-0.03, 0.00) | -0.01  (-0.02, 0.00) | -0.03  (-0.07, -0.00) | 0.01  (-0.01, 0.02) | 0.01  (-0.03, 0.04) | -0.00  (-0.00, 0.00) |
| PFOS | Baseline | 15613.1 | -0.01  (-0.01, -0.00) | NA | NA | NA | NA | NA | NA |
|  | Fiber Sources | 15630.42 | NA | -0.03  (-0.05, -0.01) | -0.01  (-0.03, 0.01) | -0.03  (-0.08, 0.01) | -0.01  (-0.02, 0.00) | -0.02  (-0.05, 0.01) | -0.01  (-0.01, -0.00) |
| PFNA | Baseline | 14523.56 | -0.01  (-0.01, -0.00) | NA | NA | NA | NA | NA | NA |
|  | Fiber Sources | 14516.11 | NA | -0.03  (-0.06, -0.01) | 0.00  (-0.01, 0.02) | -0.05  (-0.11, 0.01) | -0.03  (-0.05, -0.01) | 0.02  (-0.05, 0.09) | -0.01  (-0.01, -0.00) |

^a^ Adjustment factors are the same as for the final model in Table 3, except that total energy intake was also included as a covariate (n=6,482).

^b^ Includes grain foods, grain-based mixed dishes, pizza, and sandwiches. Does not include grain-based snacks & sweets.

Table S11. Percent change in PFAS level per inter-quartile range (IQR) increase in energy-adjusted fiber intake using multiply imputed data.^†^

|  | Percent Change (95% CI) | | |
| --- | --- | --- | --- |
| PFAS (ng/mL) | Crude Model | Full Model | Final model |
| PFOA | -5.52 (-8.19, -2.78) | -3.93 (-6.35, -1.44) | -3.69 (-6.15, -1.17) |
| PFOS | -7.67 (-10.99, -4.23) | -7.24 (-9.98, -4.41) | -7.23 (-10.02, -4.37) |
| PFNA | -6.04 (-9.03, -2.96) | -7.72 (-10.23, -5.14) | -7.98 (-10.48, -5.41) |

^†^The full model was adjusted for energy intake, consumption of meat and meat products, milk and dairy products, eggs, popcorn, fish and shellfish, age, BMI, poverty to income ratio, education, parity (in women), sex, NHANES wave, smoking, alcohol, and race/ethnicity. The final model was based on the set of adjustment variables which gave a % change in PFAS that was < 5% different from that in the full model. For PFOA, energy intake, dairy, eggs, income-to-poverty ratio and smoking were removed. For PFOS, energy intake, meat, dairy, eggs, popcorn, income-to-poverty ratio, parity, wave^2^ and alcohol were removed compared to the full model. For PFNA, energy intake, dairy, eggs, popcorn, income-to-poverty ratio, parity and alcohol were removed compared to the full model.

Table S12. Comparison of different methods of adjusting fiber intake for energy intake in models of PFAS and fiber intake (g/d) ^†^

|  |  |  | Percent Change (95% CI) | |
| --- | --- | --- | --- | --- |
| Type of energy Adjustment | Type of fiber variable | PFAS (ng/mL) | Full | Final |
| Energy used to calculate energy-adjusted fiber residual and energy intake included in model | Energy-adjusted fiber residual | PFOA | -0.47 (-0.78, -0.15) |  |
|  |  | PFOS | -0.84 (-1.21, -0.47) |  |
|  |  | PFNA | -1.48 (-2.17, -0.79) |  |
| Energy as a covariate in model | Crude fiber | PFOA | -0.48 (-0.74, -0.22) |  |
|  |  | PFOS | -0.81 (-1.14, -0.49) |  |
|  |  | PFNA | -1.10 (-1.71, -0.48) |  |
| Energy used to calculate energy-adjusted fiber residual | Energy-adjusted fiber residual | PFOA |  | -0.45 (-0.77, -0.13) |
|  |  | PFOS |  | -0.84 (-1.21, -0.46) |
|  |  | PFNA |  | -1.55 (-2.27, -0.82) |

^†^The full model was adjusted for energy intake, consumption of meat and meat products, milk and dairy products, eggs, popcorn, fish and shellfish, age, BMI, poverty to income ratio, education, parity (in women), sex, NHANES wave, smoking, alcohol, and race/ethnicity. The final model was based on the set of adjustment variables which gave a % change in PFAS that was < 5% different from that in the full model. For PFOA, energy intake, dairy, eggs, income-to-poverty ratio and smoking were removed. For PFOS, energy intake, meat, dairy, eggs, popcorn, income-to-poverty ratio, parity, wave^2^ and alcohol were removed compared to the full model. For PFNA, energy intake, dairy, eggs, popcorn, income-to-poverty ratio, parity and alcohol were removed compared to the full model.

Comment

Note that the units for energy-adjusted fiber and crude fiber are both g/d in these models. According to authoritative descriptions of the effect of using energy-adjusted residuals to represent fiber intake in a model adjusted for energy intake, when the results are compared to those from a model with crude fiber with adjustment for energy intake as a separate covariate, the beta coefficients should be identical (Willett et al. 1997; Kipnis et al. 1997). In our case, however, the results differ slightly, because when we calculated the energy-adjusted fiber values, both fiber and energy were log_e_ transformed before the regression was done, and then we back-transformed the residuals before examining the relation with PFAS. A less important reason for the difference is that in our final models, which were based on energy-adjusted residuals, energy intake as a separate variable was no longer an important confounder and was thus not included as a covariate.

Kipnis, V., Freedman, L. S., Brown, C. C., Hartman, A. M., Schatzkin, A., Wacholder, S. Effect of Measurement Error on Energy-Adjustment Models in Nutritional Epidemiology. American Journal of Epidemiology 146 (10), 842-855.

Willett, W.C., Howe, G.R., Kushi, L.H., 1997. Adjustment for total energy intake in epidemiologic studies. The American journal of clinical nutrition 65, 1220S-1228S.
